# Quality of postnatal care for mothers and neonates in Mexico: insights from the maternal eCohort study

**DOI:** 10.1101/2025.04.04.25325240

**Authors:** Martín Paredes-Cruz, Diana Perez-Moran, Svetlana V. Doubova, Catherine Arsenault, Claudio Quinzaños-Fresnedo

## Abstract

**Objective:** The study aimed to evaluate healthcare use during the postnatal period (PNC) for mothers and their babies, the content of care received, mothers’ perceived quality of care, and the factors influencing these perceptions.

**Methods:** The study analyzed postnatal survey from the maternal eCohort conducted at the Mexican Institute of Social Security (IMSS), which included 973 women aged 18 to 49 recruited after their first antenatal care visit with a family physician at 48 family medicine clinics in eight states in Mexico. We assessed PNC use, content of care and perceived quality using descriptive analysis. We used Poisson multivariable regression analysis to investigate the factors influencing women’s perceptions of higher quality of care during PNC.

**Results:** 29.4% of women and 12.0% of infants lacked healthcare during the postnatal period. Most women who received PNC attended consultations at IMSS (72.3%), while 17.7% combined IMSS with other providers, and 10% used only private services. Infants received 82.4% of recommended care, compared to 66.7% for mothers. The median perceived quality of care among women was 25 points on a scale of 8 to 40. Areas of opportunity include promoting the importance of postnatal consultations among health personnel and women, reducing waiting times, and improving the content and length of consultations. Factors associated with better perceived quality included being over 35, receiving better content of care for infants, and being treated by private providers, while lower education levels, prior pregnancies, and poor health were associated with lower perceived quality.

**Conclusion:** Improvements are needed to ensure all women and infants receive comprehensive postnatal care and perceive it as high quality.

## Introduction

Postnatal care (PNC) is an important part of the health care continuum for mothers, newborns, and children. It should prioritize prevention, timely diagnosis, and treatment of childbirth-related complications to avert morbidity and mortality while ensuring the well-being of mother-child pairs by providing evidence-based, person-centered guidance for their health care [1]. This is why the World Health Organization (WHO) and the Pan American Health Organization (PAHO) recommend examination of women and their newborns 24 hours after birth and subsequent postnatal check-ups by health professionals at least three times within the first six weeks after childbirth [1].

Despite the essential role of PNC worldwide, its median coverage rates reveal concerning gaps: 71% for mothers and 64% for infants [2], with only 41.4% of women receiving high-quality maternal PNC and 42.3% reported the same for neonatal PNC [3]. However, the existing information regarding the quality of PNC comes primarily from Africa, Southeast Asia, the Eastern Mediterranean, and Europe, with no data available on the PNC quality in Latin American countries [3].

The Mexican Institute of Social Security is recognized as the largest healthcare provider in Mexico and Latin America, serving over 72 million people, primarily workers from the formal labor sector and their families. In 2024, over 300,000 mothers delivered their babies to the IMSS hospitals. Despite the high number of births, IMSS lacks official data on the PNC usage and quality. Limited studies on this topic have revealed that only around half of mothers seek PNC, primarily due to a low perception of the need for this care [4, 5], yet there is no information on the quality of postnatal care at IMSS.

Therefore, the objective of the present study was to assess the use of PNC by mothers and their babies, the content of care they received, the quality of care perceived by mothers, and to examine the factors associated with women’s perceptions of PNC quality.

## Methods

The study analyzed data on postnatal care from the IMSS maternal eCohort [6]. The cohort consisted of 1,390 pregnant women aged 18–49 years, who were enrolled after receiving their first prenatal care consultation with a family physician at 48 IMSS family medicine clinics in eight states in Mexico. Participant recruitment took place from August to December 2023, followed by subsequent follow-up until August 2024. Detailed information on the cohort selection criteria, sample size, and sampling strategy have been previously published.

Study data were collected using a postnatal care survey questionnaire designed by the Quality Evidence for Health System Transformation (QuEST) network to assess the quality of care from the perspective of women during their postnatal care, through a telephone interview. Prior to data collection, the questionnaire was adapted and validated by the IMSS experts. The postnatal interviews were applied between 6 and 8 weeks postpartum, based on women time availability. In the present study, we defined the postnatal period as the time frame from the women’s discharge from the hospital after birth to 6-8 weeks after that.

### Study variables

To describe the study population, we collected information on women’s general characteristics including sociodemographic variables such as age, education level, marital status; paid job, primiparous mother and region of residence, comprising West (Aguascalientes and Jalisco), North (Coahuila and Nuevo León), Southeast (Veracruz and Yucatán), and Center (State of Mexico and Mexico City) regions.

Covariates on infants’ and women’s health status included: the mother’s perception of the child’s health (excellent, very good, good, regular and poor); infant health problems after hospital discharge and up until the time of the survey (i.e., feeding, bowel movement and sleep problems, as well as skin alterations) and warning signs (i.e., presence of disease with a cough, trouble breathing, fever (body temperature > 37.5 °C), experience of diarrhea with blood in the stools, hypothermia, seizures, jaundice), current child feeding (exclusive breastfeeding, formula, both). The current women health status was measured using self-rated health (excellent, very good, good, regular and poor) and the risk of postnatal depression that was evaluated with the Patient Health Questionnaire (PHQ-9) validated in Mexico [7]. This scale has nine questions and four response options ranging from 0 (none of the days) to 3 (almost every day), with a grand total between 0 and 27. The severity of the symptoms is organized into six categories of risk of depression: 0-4 (minimal), 5-9 (mild), 10-14 (moderate), 15-19 (moderate to severe) and 20-27 (severe). For the purposes of this research and considering the low prevalence of women who scored ≥10 on the PHQ-9, we defined the risk of depression as women who scored ≥5 during the postnatal period.

### PNC utilization

PNC utilization was assessed by a categorical variable for the number and type of medical consultations by mother-infant pairs (none, mothers only, infants only, or both mothers and babies); the number of consultations for mothers and their infants; postnatal check-ups for mothers and babies or other types of consultations with health professionals during this period (i.e., consultations related to health problems, neonatal screening, vaccinations, or other concerns); the type of healthcare providers used (such as IMSS or other providers like private provider or the Ministry of Health), or use of both IMSS and other providers; and the reasons for not receiving care in the postnatal period.

### Content of PNC for infants

We surveyed women on clinical activities performed by health providers for the baby (measuring weight, height, head, chest, abdomen and body temperature, listening to the chest with a stethoscope, examination of eyes, hearing, reflexes, genitals and hip bones), advices on baby care including vaccines that the baby should receive, warning signs that require medical care, breastfeeding and its frequency, safe sleeping positions, cases that require follow-up with a pediatrician or a neonatologist and how to play and interact with the baby. We also generated a variable that describes the percentage of activities provided to the baby regarding required PNC clinical activities, including all previously defined activities, with a minimum score of 0% and maximum of 100%. In addition, we collected information on the health provider’s recommendations regarding baby’s warning signs and current baby’s vaccination status regarding BCG vaccine, pentavalent vaccine, pneumococcal vaccine and rotavirus vaccine.

### Content of PNC for women

During the postnatal period, we assessed the clinical activities performed for the mothers, including blood pressure and body temperature measurements, C-section scar examination (for women who had a c-section), advice on women’s warning signs during postnatal period, mood, breast care, physical activity, as well as on reinitiating sexual activity after childbirth and family planning options. Furthermore, we generated a variable that describes the percentage of activities provided to each woman regarding required PNC clinical activities, including all previously defined activities.

### Dependent variable Perceived quality

Women’s perception about quality of care during medical consultations in the postnatal period was measured by asking them to rate eight components of their care on a scale of 1 to 5 (poor, fair, good, very good, and excellent): (i) the level of respect shown by the physician; (ii) courtesy and helpfulness of non-medical staff; (iii) the extent to which the physician involved women in decisions about their care or care of their child; (iv) availability of medical equipment or access to laboratory tests; (v) clarity of the physician’s explanations; (vi) knowledge and skills of the physician; (vii) the amount of time the physician spent with the woman; and (viii) waiting time [6]. We calculated a summary score of perceived PNC quality as the sum of the response scores for these eight indicators, with a minimum score of 8 and maximum of 40 points [6].

### Sample size

Detailed information regarding the cohort’s sample size, and sampling strategy have been published previously [6]. However, out of 1,390 women who agreed to participate in the maternal eCohort in Mexico and completed the baseline questionnaire, only 973 (70%) answered the postnatal care survey. Among the rest, 3.7% (n=51) experienced a miscarriage, 0.6% had stillbirths (n=5) or neonatal deaths (n=3), and 25.7% (n=358) dropped out. The sample of 973 participants is sufficient to address the present study objectives, as it allows meeting the assumption of ensuring at least ten participants per covariate included in the multivariable regression analysis [8].

### Statistical analysis

We used descriptive statistics to describe women and infants’ health and demographic characteristics, PNC utilization, content of care, and perceived quality. We used percentages for categorical variables, median with range (minimum and maximum values) for non-normally distributed numerical variables, verified by the Shapiro-Wilk test.

Second, to analyze the factors associated with perceived PNC quality of care, we performed a multivariable Poisson regression analysis. We used Poisson regression, taking into account that our dependent variable (perceived quality of care score) is a count variable measured on the scale that ranges from 8 points (poor quality) to 40 points (excellent quality), approaching to the Poisson distribution and because this regression is recommended for analyzing numerical dependent variables that represent counts of events distributed in a way that follows a Poisson distribution. Our modeling strategy was based on the VanderWeele and Shpitser criteria for the selection of confounders. These authors recommend including all conceptual and clinically relevant covariates to ensure that the final model adjusts for even mild confounders. In addition, the standard errors of the regression model were adjusted for clustering based on the women’s region of residence. The multivariable analysis on perceived PNC quality focused solely on women who had at least one consultation for themselves or their baby during the postnatal period (n=901); however, because 16 participants (1.8%) had missing data for one or more study variables, they were automatically excluded from the analysis resulting in an analytic sample of 885 participants. Moreover, for the analysis of infants outcomes, we analyzed the information of one infant per woman because of the limited number of twins in the cohort (n=12), and since the outcomes for the twins were similar, we only provided details about the firstborn twin.

A p-value of ≤0.05 was considered statistically significant. We analyzed the data using Stata 14 statistical software (Stata Corp LP; College Station, TX). The study is reported according to the STROBE (Strengthening the Reporting of Observational Studies in Epidemiology) guidelines.

### Ethics

The study was approved by the IMSS National Research and Ethics Committees (R-2022-785-064). Before participating in the study, all women provided written informed consent form.

## Results

The majority of survey participants (86.1%) were 34 or younger, mostly married (84.7%), and had high school education (64.5%). More than half (60.4%) had remunerated jobs, and 36.5% were primiparous. Most participants were from the north (30.2%), followed by the central (25.9%), west (22.8%), and south (21.1%) (Table 1).

**Table 1.**
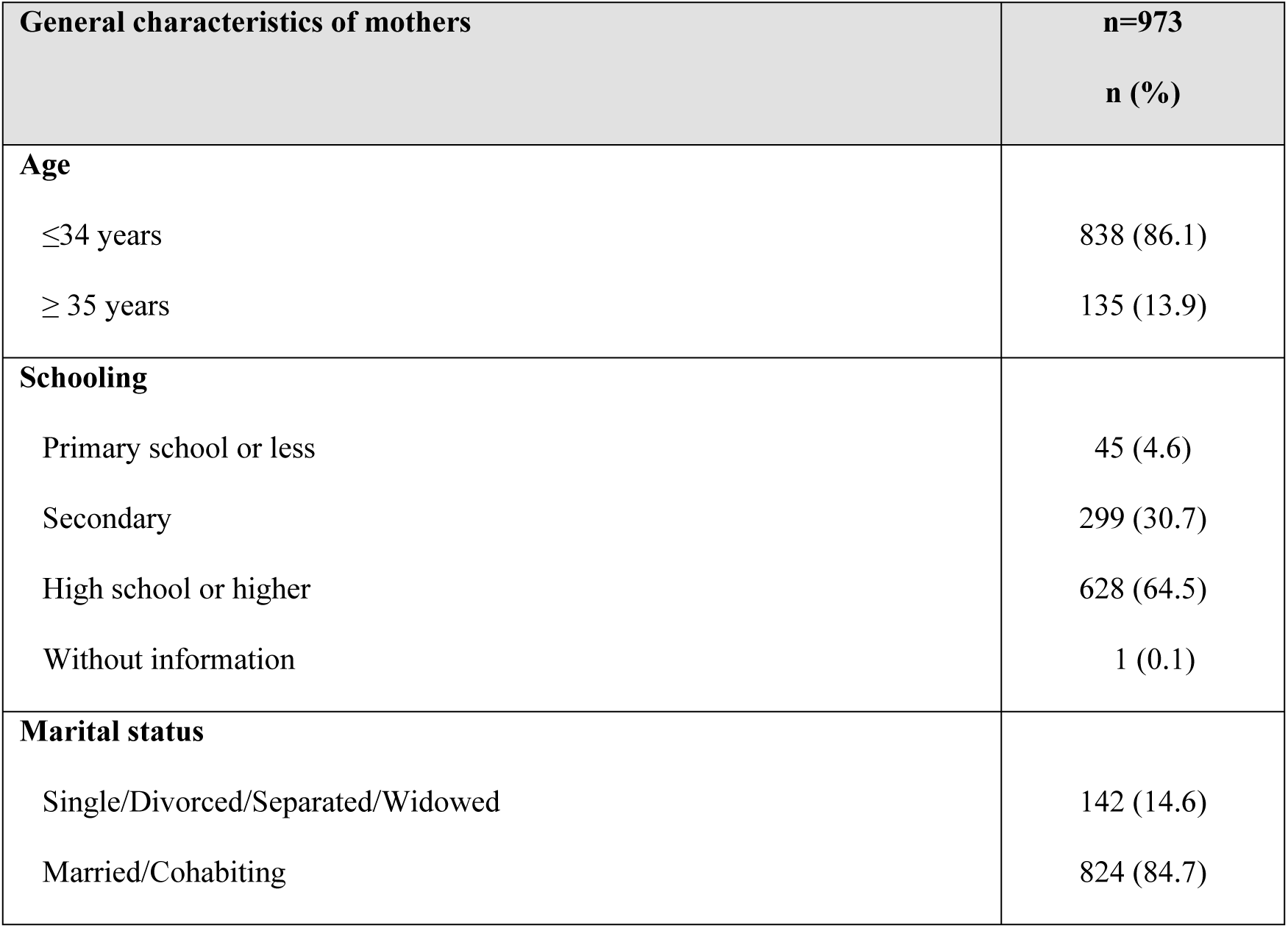
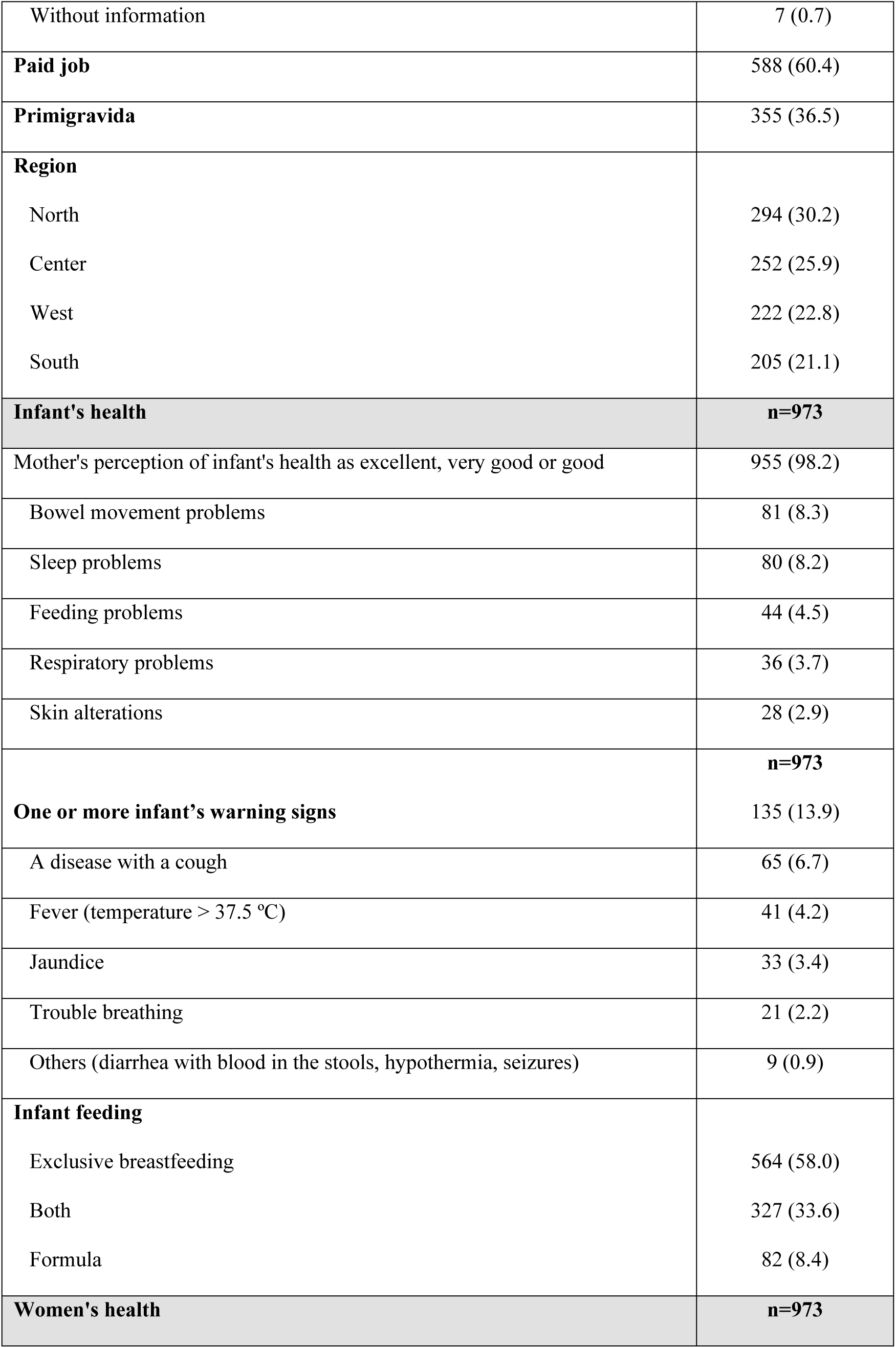
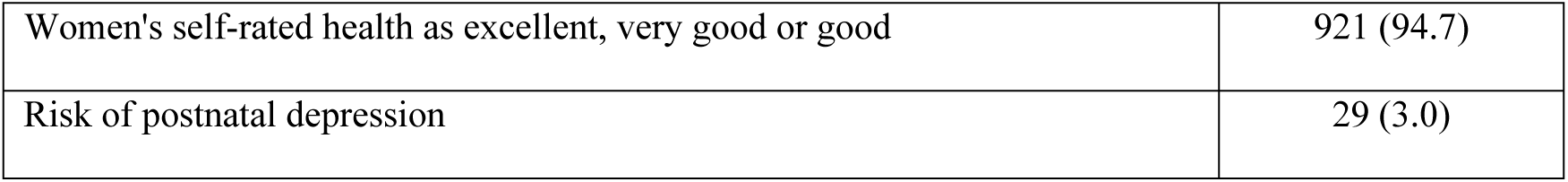
General characteristics and health of mothers and infants.

In the postnatal period, 98.2% reported their babies were in excellent or good health, with fewer than 10% facing some health issues such as bowel movement problems (8.3%), sleep difficulties (8.2%), feeding challenges (4.5%), respiratory issues (3.7%), or skin disorders (2.9%). In addition, 13.9% of babies exhibited warning signs, primarily cough (6.7%) and fever (4.2%). Feeding practices showed that 58% provided exclusive breastfeeding, 33.6% combined breastfeeding with formula, and 8.4% used formula alone. Most women (94.7%) rated their health as excellent or good. Only 3% presented a risk of postpartum depression (Table 1).

7.4% of participants did not seek medical consultations after being discharged from the hospital following childbirth. Of those who had consultation with healthcare provider during the postnatal period, 4.6% were solely for women, 22% exclusively for infants, and 66% attended consultations for both mothers and their babies.

Among infants, 83.7% attended at least one PNC visit, compared to 67.4% of mothers. One third (34.6%) accessed additional consultations, mainly for neonatal screening (46.9%), vaccinations (43.9%), or illnesses (19.3% for children and 11.0% for mothers) (Table 2). Most received postnatal care at IMSS (72.3%), while 17.7% used both IMSS and private services, and 10% only private healthcare providers. The primary reason for not seeking PNC was the belief that neither they nor their baby needed a consultation (40.3%), followed by appointment unavailability (19.4%) and long waiting times (12.5%) (Table 2).

**Table 2.**
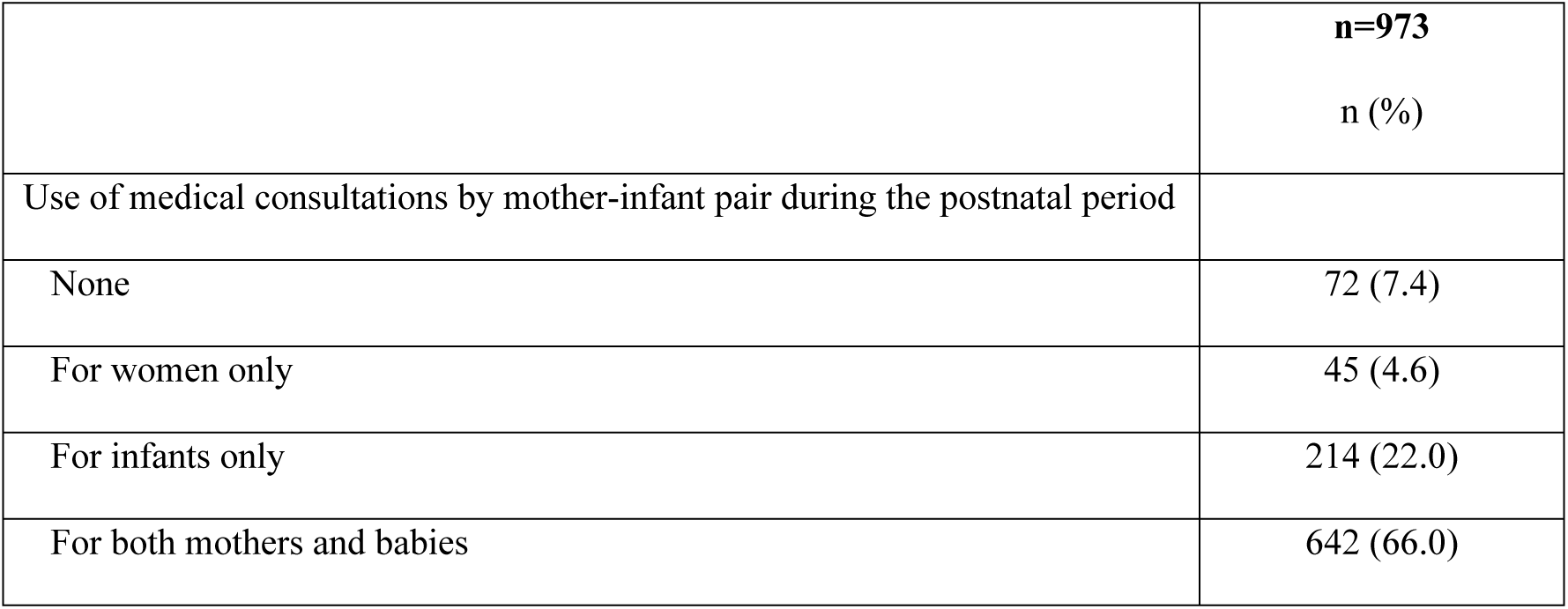
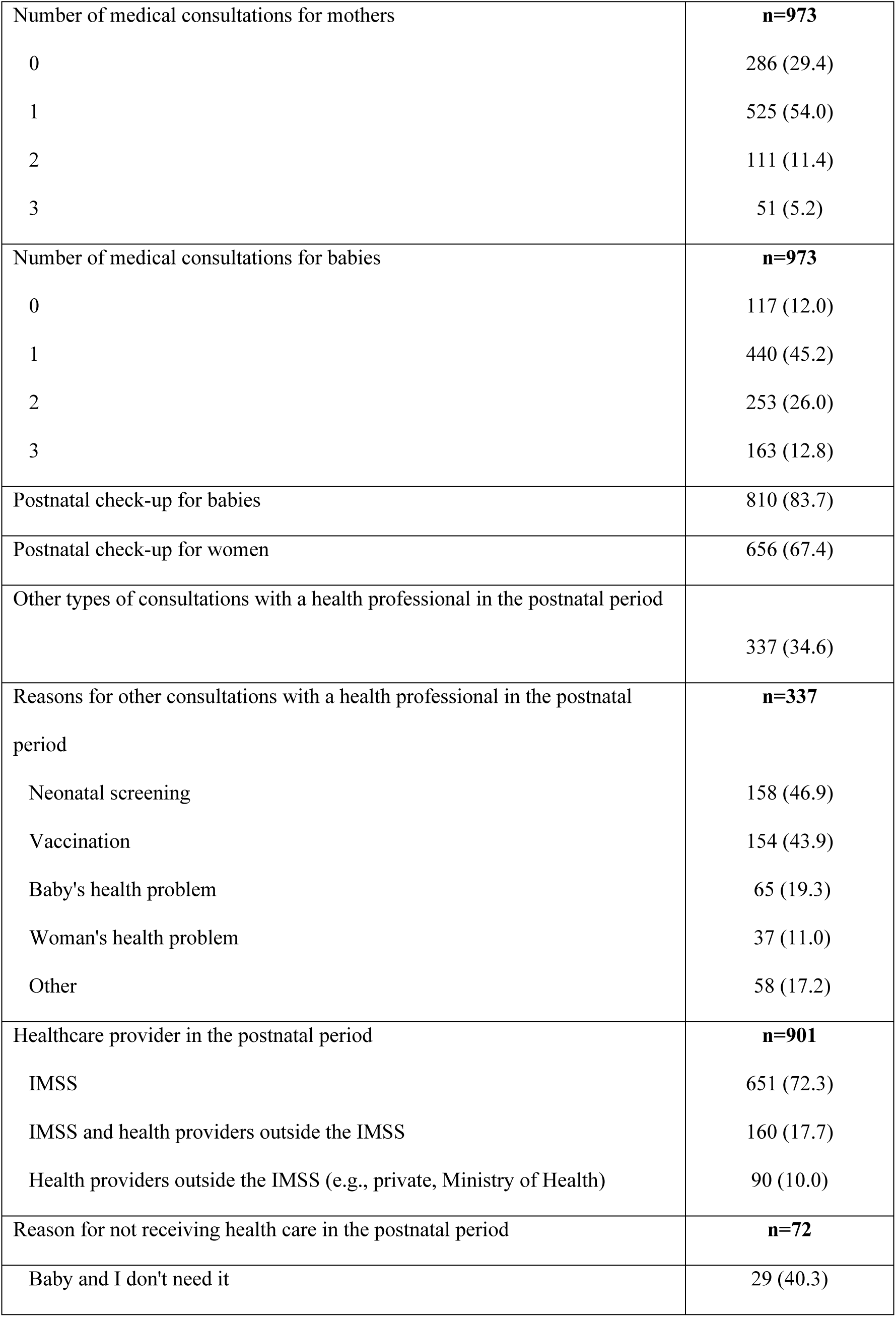
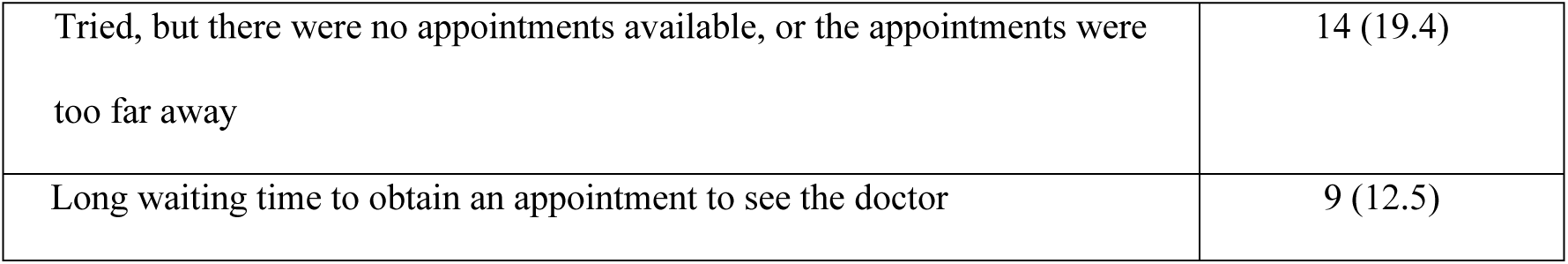
Healthcare use during the first 6 to 8 weeks after childbirth and discharge from the hospital.

During child postnatal care, over 94% of women reported having their babies’ weight, height, and temperature measured. Additionally, 92.2% had their babies’ chests examined with a stethoscope, while 81.3% had their eyes checked and 81.0% had their tendon reflexes assessed. At least 71.6% of babies had their genitals, hearing, and hip bones examined. Furthermore, most mothers received guidance on vaccinations (91.8%), warning signs for medical attention (85.8%), breastfeeding (81.9%), feeding frequency (74.4%), and safe sleeping positions (68.1%). Only 54.8% were advised on follow-up visits with a pediatrician, and 51.5% received advice on interacting with their babies. The median percentage of clinical activities provided to babies was 82.4%, ranging from 0 to 100% (Table 3).

**Table 3.**
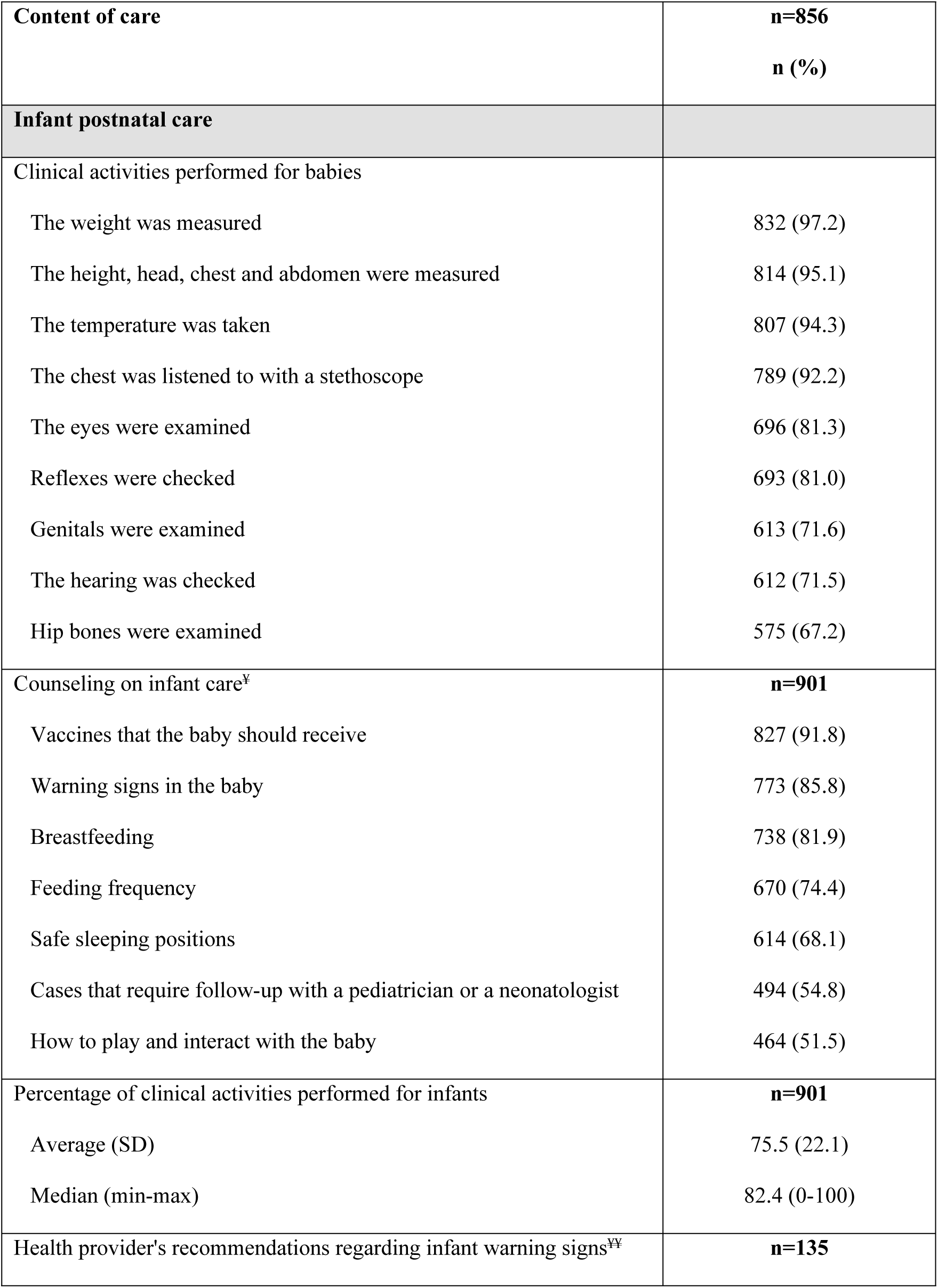
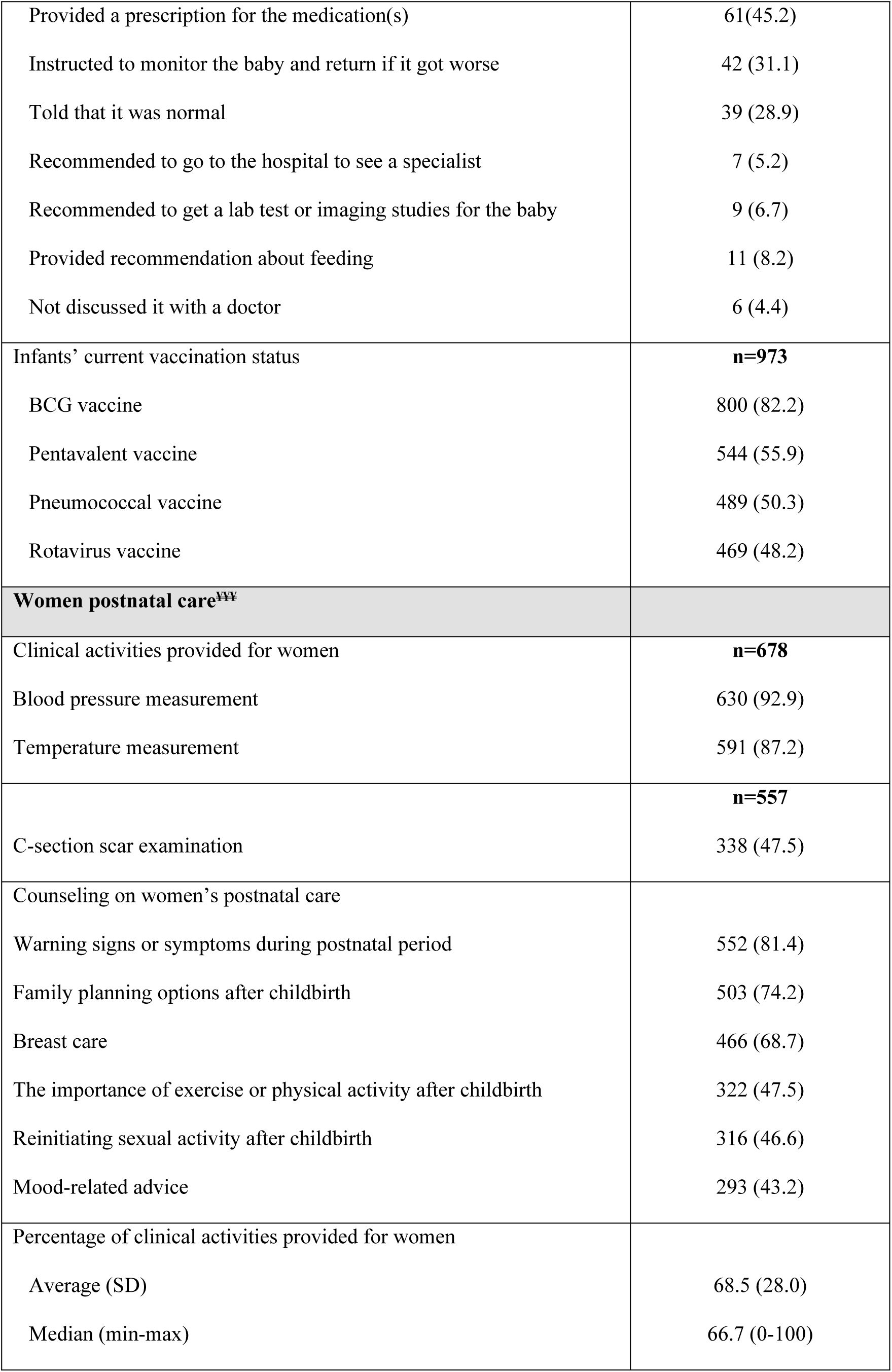
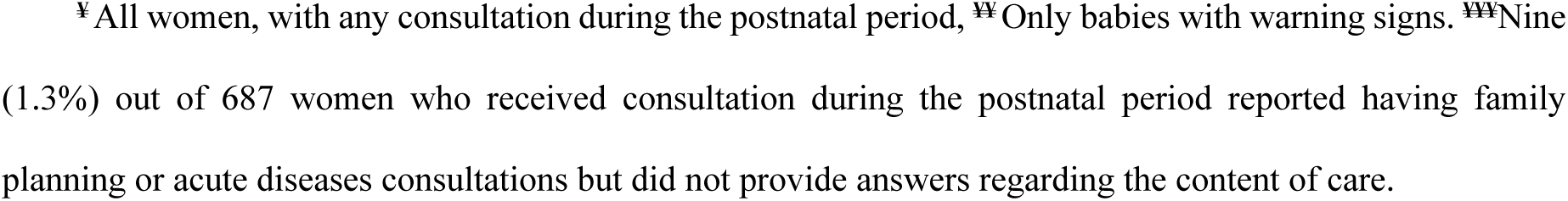
Content of infant and women postnatal care in the first 6 to 8 weeks after childbirth.

| **Content of care** | **n=856**<br>**n (%)** |
| --- | --- |
| **Infant postnatal care** |  |
| Clinical activities performed for babies |  |
| The weight was measured | 832 (97.2) |
| The height, head, chest and abdomen were measured | 814 (95.1) |
| The temperature was taken | 807 (94.3) |
| The chest was listened to with a stethoscope | 789 (92.2) |
| The eyes were examined | 696 (81.3) |
| Reflexes were checked | 693 (81.0) |
| Genitals were examined | 613 (71.6) |
| The hearing was checked | 612 (71.5) |
| Hip bones were examined | 575 (67.2) |
| Counseling on infant care<sup>xy</sup> | **n=901** |
| Vaccines that the baby should receive | 827 (91.8) |
| Warning signs in the baby | 773 (85.8) |
| Breastfeeding | 738 (81.9) |
| Feeding frequency | 670 (74.4) |
| Safe sleeping positions | 614 (68.1) |
| Cases that require follow-up with a pediatrician or a neonatologist | 494 (54.8) |
| How to play and interact with the baby | 464 (51.5) |
| Percentage of clinical activities performed for infants | **n=901** |
| Average (SD) | 75.5 (22.1) |
| Median (min-max) | 82.4 (0-100) |
| Health provider's recommendations regarding infant warning signs<sup>yy</sup> | **n=135** |
| Provided a prescription for the medication(s) | 61(45.2) |
| Instructed to monitor the baby and return if it got worse | 42 (31.1) |
| Told that it was normal | 39 (28.9) |
| Recommended to go to the hospital to see a specialist | 7 (5.2) |
| Recommended to get a lab test or imaging studies for the baby | 9 (6.7) |
| Provided recommendation about feeding | 11 (8.2) |
| Not discussed it with a doctor | 6 (4.4) |
| Infants' current vaccination status | **n=973** |
| BCG vaccine | 800 (82.2) |
| Pentavalent vaccine | 544 (55.9) |
| Pneumococcal vaccine | 489 (50.3) |
| Rotavirus vaccine | 469 (48.2) |
| **Women postnatal care<sup>xxx</sup>** |  |
| Clinical activities provided for women | **n=678** |
| Blood pressure measurement | 630 (92.9) |
| Temperature measurement | 591 (87.2) |
|  | **n=557** |
| C-section scar examination | 338 (47.5) |
| Counseling on women's postnatal care |  |
| Warning signs or symptoms during postnatal period | 552 (81.4) |
| Family planning options after childbirth | 503 (74.2) |
| Breast care | 466 (68.7) |
| The importance of exercise or physical activity after childbirth | 322 (47.5) |
| Reinitiating sexual activity after childbirth | 316 (46.6) |
| Mood-related advice | 293 (43.2) |
| Percentage of clinical activities provided for women |  |
| Average (SD) | 68.5 (28.0) |
| Median (min-max) | 66.7 (0-100) |

<sup>¥</sup>All women, with any consultation during the postnatal period, <sup>₩</sup>Only babies with warning signs. <sup>₩₩</sup>Nine (1.3%) out of 687 women who received consultation during the postnatal period reported having family planning or acute diseases consultations but did not provide answers regarding the content of care.

Regarding the current baby’s vaccination status, 82.2% received the BCG vaccine, 55.9% the pentavalent vaccine, 50.3% the pneumococcal vaccine and only 48.2% the rotavirus vaccine (Table 3).

70.6% of women had medical consultation in postnatal period. During postnatal care for women, the majority had their blood pressure (92.9%), and temperature (87.2%) measured. Among those with cesarean sections, only 47.5% had their surgical scars examined. Additionally, 81.4% received advice on warning signs during the postnatal period, 74.2% on family planning, and 68.7% on breast care. However, fewer than half were advised on exercise (47.5%), reinitiating sexual activity (46.6%), and mood status (43.2%). The median percentage of clinical activities performed for women was 66.7%, ranging from 0% to 100% (see Table 3).

When asked about their perception of the quality of care during the postnatal period, 95.3% of women rated the level of respect shown by their physician as excellent, very good, or good; 93.5% rated the courtesy and kindness of the staff at the family medicine clinic in the same favorable categories. Furthermore, 92.2% of women felt that the physician effectively involved them in decisions regarding their care, while 92% rated the equipment and materials available for consultations as excellent, very good, or good. The clarity of the physician’s explanations and the physician’s knowledge and skills were rated as excellent, very good, or good by 91% of women. In terms of the time the physician spent during the consultation, 88.8% rated it as excellent, very good, or good, while the waiting time was rated in the same way by only 77.9% (Fig 1). Overall, the median score for the quality of care perceived by women was 25 points, with scores ranging between 10 and 40 points.

**Fig 1.**
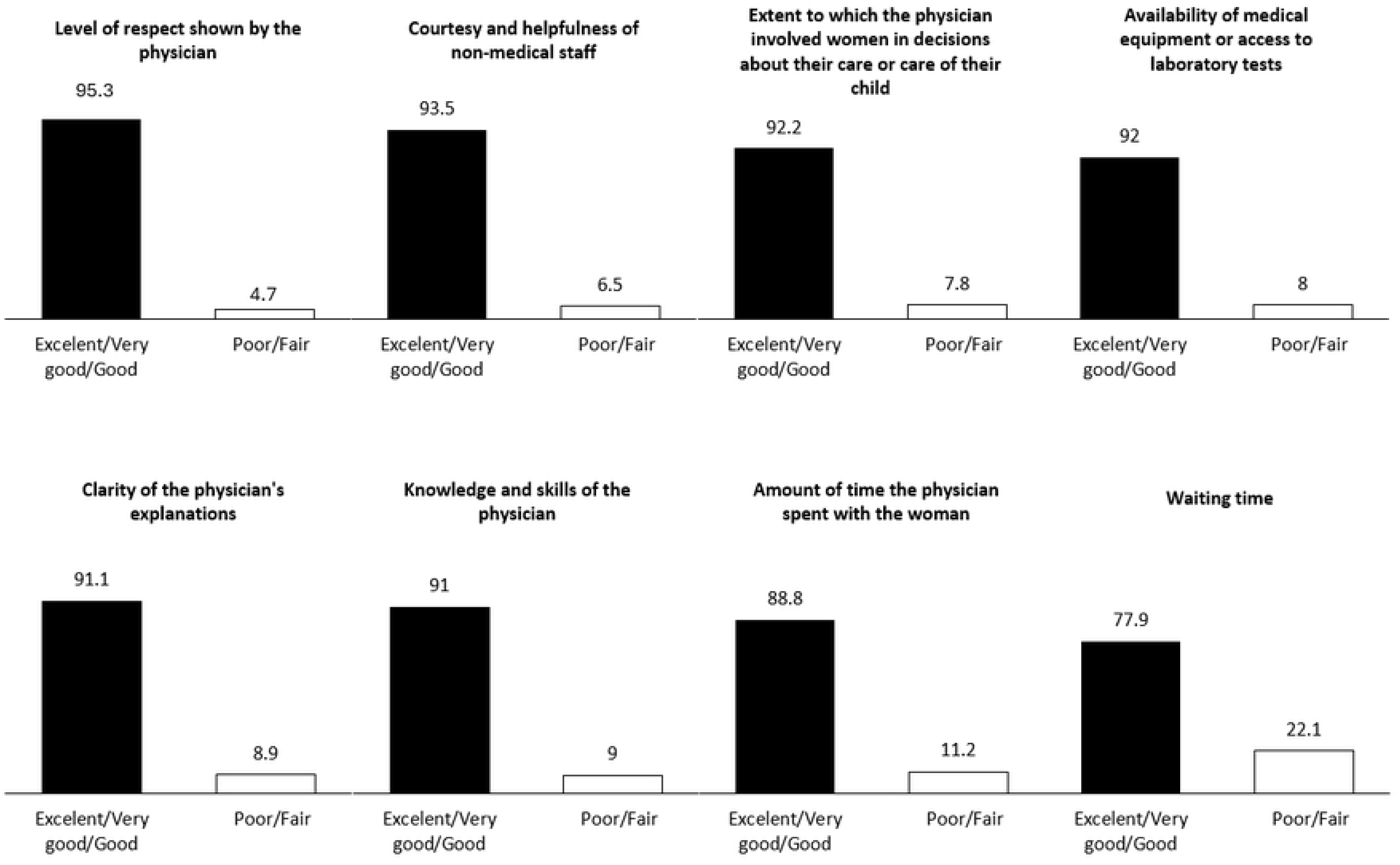
Perception of quality of postnatal care.

Table 4 presents the factors associated with women’s perceptions of care quality during the postnatal period. Notably, women aged 35 years and older, as well as those who received a higher number of clinical activities for babies, and those who sought care outside of the IMSS, reported a better perception of care quality. In contrast, women with a lower level of education, those with previous pregnancies, and those who perceived the health of the baby and mother as poor, tended to have a lower perception of the quality of care they received.

**Table 4.** Results of Poisson multivariable regression for the factors associated with perceived quality of postnatal care, n=885.

| Variable | Adjusted prevalence ratio | Robust standard errors | 95% CI | p |
| --- | --- | --- | --- | --- |
| **General characteristics of mothers** |  |  |  |  |
| Age ≥ 35 years | **1.06** | **0.03** | **1.002, 1.123** | **0.042** |
| Schooling |  |  |  |  |
| Primary school or less | **0.93** | **0.02** | **0.89, 0.97** | **0.001** |
| Secondary school | **0.98** | **0.005** | **0.97, 0.99** | **<0.001** |
| Marital status |  |  |  |  |
| Single/Divorced/Separated/Widowed | 1.01 | 0.03 | 0.96, 1.07 | 0.665 |
| Remunerated job | 0.99 | 0.01 | 0.97, 1.02 | 0.727 |
| With previous pregnancies | **0.98** | **0.002** | **0.97, 0.98** | **<0.001** |
| **Current health status** |  |  |  |  |
| Mother's perception of infant's health as regular or poor | **0.91** | **0.03** | **0.85, 0.98** | **0.013** |
| One or more child's warning signs | 0.99 | 0.03 | 0.95, 1.05 | 0.921 |
| Women's self-rated health as regular or poor | **0.87** | **0.03** | **0.81, 0.93** | **<0.001** |
| Risk of postnatal depression | 1.02 | 0.06 | 0.92, 1.14 | 0.677 |
| **Healthcare during the postnatal period** |  |  |  |  |
| Percentage of clinical activities provided for babies | **1.003** | **0.00006** | **1.0033, 1.0035** | **<0.001** |
| Percentage of clinical activities provided for women | **1.0005** | **0.0003** | **0.999, 1.001** | **0.055** |
| Healthcare provider outside the IMSS | **1.16** | **0.08** | **1.02, 1.32** | **0.026** |
| IMSS and other health care providers | 1.002 | 0.04 | 0.92, 1.09 | 0.955 |

Dependent variable: perceived quality of care by women during postnatal period. Reference categories: <35 years; high school or higher; unemployed; primipara; mother's perception of infant's health as excellent, very good or good, no infants warning signs; women's self-rated health as excellent, very, good or good; without risk of postnatal depression; IMSS health care provider.

## Discussion

The present study used data from the maternal eCohort that followed 973 women from their first ANC visit in an IMSS family medicine clinic until 6 to 8 weeks postpartum. We found that one-third of women and 12% of infants received no PNC after post-delivery hospital discharge. Most mothers and infants attended PNC at IMSS, while 17.7% combined IMSS services with private care, and the remaining 10% exclusively utilized private or other health services apart from the IMSS. During PNC visits, infants received 82.4% of recommended care, compared to 66.7% for mothers. Most poorly rated aspect of care included wait time and time spent with the provider during a consultation. The median perceived quality of care among women was 25 points on a scale of 8 to 40. Factors associated with better perceived PNC quality included being over 35, receiving better content of care for infants, and being treated by private providers, while lower education levels, prior pregnancies, and poor health were associated with lower perceived quality.

Postnatal care services are necessary for achieving Sustainable Development Goals related to reproductive health, particularly in reducing maternal and neonatal mortality rates [1]. Yet, despite their importance, postnatal care remains the least prioritized intervention within the continuum of maternal and childcare [9]. The study found that one-third of women and 12% of infants had no medical consultations during the postnatal period (until 6 to 8 weeks) after being discharged from the hospital following childbirth. The maximum number of visits recorded for mothers and babies was three, consistent with WHO recommendations for at least three postnatal visits after discharge (within 24 hours for vaginal deliveries and 72 hours post-caesarean section), and within 6-8 weeks following childbirth. Using data from 48 LMICs (4 in the Americas, 13 in Asia, and 31 in Africa), The Countdown to 2030 Collaboration found that 59% of mothers and 42% of infants received postnatal care [10]. Using data from 74 LMICs (1 in Europe, 2 in Oceania, 6 in the Americas, 18 in Asia, and 47 in Africa), Victora et al found that 58% of mothers received PNC, while only 28% of infants did [11]. In our study, PNC coverage was 67% for mothers and 84% for infants. There are many reasons for higher rates of PNC use in our study including the fact that Mexico is a wealthier country that those included in the Countdown and Victora studies, the data is more recent, and the women enrolled in the IMSS Maternal eCohort were recruited in health facilities (and hence received ANC) rather than being recruited in the population. These poor levels of PNC use for mothers and their infants are alarming, especially considering WHO recommendation for integrated postnatal care for both the mother and infants. The Amouzou et al. suggest these gaps may be linked to poor quality of postnatal care, insufficient understanding of postnatal care benefits, and the separation of mothers and newborns after birth, resulting in only one receiving postnatal care [12]. In our study, the main reasons for not participating in postnatal consultations were the belief that neither the mother nor the infant required health care (40.3%), followed by difficulties in obtaining an appointment (19.4%), and important waiting times (12.5%). These findings are similar to those reported by Bancalari in 2022 in Mexico [5],^5^ Zamawe in 2015 in Malawi [13], and Tesfahun in 2014 in Ethiopia [14], where a majority of mothers indicated a low perception of the necessity to pursue postnatal care if their newborns appeared healthy or if they thought the health issue would resolve on its own. It is also in line with data from 20 countries (16 in Africa, 2 in Latin America, and 2 in Asia) [15], where the predominant reason for not attending postnatal care was attributed to long waiting times for appointments.

Technical quality, which includes disease or condition specific clinical actions, along with the respectful and supporting relationship between healthcare professionals and patients, are vital for high-quality care. The current vision of WHO and PAHO emphasizes importance of not just life-saving interventions but also the well-being of women and their infants through positive postnatal experiences and care advice. In our study, the clinical activities necessary to perform in the postnatal period were identified by the QuEST network expert group and the IMSS clinical experts considering the recommendations of WHO and PAHO. Analysis of these activities identified that those infants received 82.4% of recommended activities, while women received only 66.7%. The least performed clinical activities for infants were the physician’s recommendation on when to take the baby to the pediatrician, and how to interact with the baby, while many women were not advised about physical exercise, resuming sexual activity, or preventing postpartum depression. Currently, research primarily focuses on the first 48 hours after delivery, with limited studies investigating care during the subsequent six weeks, as outlined by the WHO. A relevant study from Ghana indicated that mothers often receive inadequate explanations about their postnatal care. In some instances, infants are separated from their mothers and taken to another room for clinical procedures, leaving mothers unaware of the activities being performed [16]. Therefore, our study provides crucial evidence regarding the clinical care that women and their babies receive in the 6-8 weeks following post-delivery discharge. We identified a significant need to promote the importance of postnatal consultations, counseling them on physical exercise, addressing the resumption of sexual activity, preventing and detecting postpartum depression, and providing guidance on childcare.

### A positive postnatal experience is an essential component of high-quality care

The WHO characterizes a positive postnatal experience as one where women, newborns, and their families are provided with consistent information, reassurance, and support from healthcare staff, and where a well-resourced and adaptable health system acknowledges the needs of women and babies while honoring their cultural backgrounds [1]. In our research, the median perceived quality of postnatal care among women was 25 points on a scale ranging from 8 to 40 points, with most women rating the respect shown by doctors and the courtesy of other healthcare staff as excellent or good, and less rating the same the waiting time and the duration of medical consultations. In earlier assessments of our cohort, we observed that the average waiting period to see a physician was 30 minutes [6]. Although waiting time is often overlooked in postnatal consultations, a qualitative study conducted in Zanzibar indicated that healthcare personnel believe patients avoid attending postnatal consultations due to extended waiting times, perceiving them as fatigued and feeling like their time is wasted [17].

Furthermore, long waiting times can be particularly frustrating for women attending with their newborns, due to the concern about exposing them to potential infections in the clinic, as highlighted in a study conducted in Ethiopia [18]. Regarding the duration of consultations it varies based on the specific needs of each woman and her newborn. Our study indicates that one in ten women rated the duration of their consultation as either regular or poor. Previous analysis of prenatal care of this cohort revealed an average consultation time of 20 minutes, with more than half of the women describing it as regular or poor [6]. Currently, there are no institutional or international guidelines recommending the duration of postnatal consultations. However, these consultations should last long enough to complete all necessary clinical activities for both the mother and her newborn. They should also provide essential information, support, and an opportunity for mothers to address any questions they may have.

Our study found that a higher percentage of clinical activities provided to both the mother and baby during the postnatal period is linked to a better perception of healthcare quality. This finding is significant as it underscores the importance of necessary clinical activities not only for the health of the mother-child binomial, but also for enhancing the perception of quality. This aspect had not been identified in previous studies on the quality of postnatal care.

We found that those over 35 years old had a higher perception of quality, echoing findings from Kenya, Nepal and Uganda, which suggest older women are more experienced in seeking and advocating for quality postnatal care [19, 20]. Additionally, using private health services was associated with a better perception of quality, corroborating studies from Mexico and Nepal, which showed that private healthcare users rated their quality of care higher than public sector users [21, 22]. This is often attributed to shorter waiting times and more user-centered care in private facilities.

In our study, being multiparous was linked to a lower perception of quality, similar to findings from Sweden [23], where researchers found that primiparous women were more satisfied with the quality of postnatal care than multiparous women.

We also found that lower education was associated with a greater likelihood of perceiving lower quality of care. This is different from previous research indicating that individuals with lower education may have lower expectations and a poorer understanding of medical procedures, leading them to be less critical of their healthcare and be more satisfied compared to those with higher education [24]. Our finding suggests the possible inequality of care for women with lower education levels. This underscores the need to improve healthcare for these women. One way to do this is by using clearer, simpler language that everyone can understand instead of complicated medical terminology and by explaining the clinical procedures provided.

Our research has several limitations. Firstly, all data used in this study were self-reported by participants, which may lead to some discrepancies between the care provided and what they remembered. Also, women with more education or prior childbirth experience may better recall or understand PNC content and the counseling received. However, differential recall by less educated women might indicate that health providers must tailor their communication style and counseling intensity to the client’s level of education and comprehension. The survey collected data on postnatal care visits within the first 6-8 weeks postpartum. Recall bias may occur, with participants likely to remember recent visits more accurately than earlier ones, though the interval between consultations and interviews was no longer than one month. Finally, participants’ responses might be influenced by social desirability bias, providing answers they think the interviewer expects; however, the telephone interviews may encourage more honest responses by excluding nonverbal cues.

## Conclusions

Improvements are needed to ensure all women and infants receive comprehensive postnatal care and perceive it as high quality. Areas of opportunity include promoting the importance of postnatal consultations among health personnel and women, reducing waiting times, and improving the content and length of consultations.

## Abbreviations

IMSS: Mexican Institute of Social Security
PAHO: Pan American Health Organization
PNC: Postnatal care
QuEST: Quality Evidence for Health System Transformation network
WHO: World Health Organization

## Authors’ contributions

Conceptualization: SVD, CA, CQF, MPC & DPM

Methodology: SVD, CA

Data Curation: MPC, SVD, DPM

Formal Analysis: SVD, MPC

Writing – original draft: SVD, MPC, DPM

Writing – Review & Editing: SVD, MPC, DPM, CA, CQF

Final approval of the version to be published: MPC, DPM, SVD, CA, CQF

## Data Availability Statement

The entire dataset is available at https://github.com/svetlanadoubova/PNC_Mex

## Acknowledgements

The authors would like to thank Ingrid Patricia Martinez Vega and Ana Maria Lira Reyes for their support in the fieldwork of the study.

